# Epidemiological model can forecast COVID-19 outbreaks from wastewater-based surveillance in rural communities

**DOI:** 10.1101/2024.02.01.24302131

**Authors:** Tyler Meadows, Erik R. Coats, Solana Narum, Eva Top, Benjamin J. Ridenhour, Thibault Stalder

## Abstract

Wastewater can play a vital role in infectious disease surveillance, especially in underserved communities where it can reduce the equity gap to larger municipalities. However, using wastewater surveillance in a predictive manner remains a challenge. We tested if detecting SARS-CoV-2 in wastewater can predict outbreaks in rural communities. Under the CDC National Wastewater Surveillance program, we monitored several rural communities in Idaho (USA). While high daily variations in wastewater viral load made real-time interpretation difficult, a SEIR model could factor out the data noise and forecast the start of the Omicron outbreak in five of the six cities that were sampled soon after SARS-CoV-2 quantities increased in wastewater. For one city, the model could predict an outbreak 11 days before reported clinical cases began to increase. An epidemiological modeling approach can transform how epidemiologists use wastewater data to provide public health guidance on infectious diseases in rural communities.

## Introduction

Wastewater-based epidemiology (WBE) is a promising approach for broad-scale, agnostic surveillance of infectious diseases and antimicrobial resistance within and across communities. Indeed, many infectious agents such as SARS-CoV-2, poliovirus, RSV, and flu shed through stool (1,2), and potentially urine (3), and are thus detectable in wastewater (4,5). Additionally, WBE assesses infectious disease circulation in both symptomatic and asymptomatic populations. Most importantly, WBE can improve and accelerate the early detection of infectious disease outbreaks by public health authorities, providing actionable data for epidemiologists. However, barriers remain to how epidemiologists might use or interpret this data to inform public health in a predictive way rather than a retrospective approach. A modeling-based approach to wastewater data can provide the framework to interpret infectious disease spread and burden by estimating epidemiological parameters such as incidence (6), prevalence (7–9), or effective reproductive number (10).

WBE also presents a unique opportunity to support vulnerable and underserved communities (11). In particular, rural communities are at a higher risk of severe outcomes associated with certain infectious diseases due to demographic factors (e.g., age), underlying healthcare challenges (e.g. obesity, smoking), limited resources (12–15), or reduced risk perception (16). Moreover, as rural communities often lack the resources for broader-scale clinical testing, WBE can help sustain rural community health. Unfortunately, WBE has mainly focused on urban areas and larger cities, leaving rural communities with reduced access to this critical data (11,17–20). In that regard, WBE is a tool that promotes equity (21).

The COVID-19 pandemic provided a unique opportunity to leverage and expand WBE while concurrently establishing greater value for epidemiologists. Concentrations of SARS-CoV-2 found in wastewater correlate well with the number of COVID-19 cases (22–24). Thus, with the right tools in place WBE can serve as an early warning for a potential COVID-19 outbreak (4,25–28). Perhaps more critically for epidemiologists, the utility of wastewater-based detection of SARS-CoV-2 has become even more significant with reduced COVID-19 clinical testing due to home tests and general lack of clinical reporting (29). However, there remains a gap in translating WBE data in a timely manner for use by public health officials. Moreover, a primary obstacle for epidemiologists is the variability and uncertainty inherent in wastewater monitoring (30,31).

Modeling SARS-CoV-2 wastewater data represents an opportunity to bridge the information gap with epidemiologists while bringing enhanced and timely health monitoring to rural communities. Here we present and discuss a susceptible-exposed-infectious-recovered (SEIR) epidemiological model developed based on wastewater detection of SARS-CoV-2 for epidemiological surveillance of COVID-19 in rural area. We specifically tested the hypothesis that the detection and quantification of SARS-CoV-2 in wastewater forecasts the start of an outbreak in rural communities.

## Material & Methods

### Sites and sample collection

Wastewater samples were collected from wastewater treatment facilities (WWTFs) located in five rural communities serving approximately 1,000 or less inhabitants and a small city in a rural county in Idaho, USA (Table 1). These rural communities are defined as "rural" according to the 2020 U.S. Census Bureau. The county itself falls under the category of a "rural" or "nonmetropolitan" county, as classified by the U.S. Office of Management and Budget. Rural cities are abbreviated RC1 to RC5 and the small city SC. All WWTFs primarily treat domestic wastewater; the SC WWTF also receives effluent from a regional hospital.

**Table 1:** Site characteristics and outbreak detection from the wastewater data and clinically confirmed cases. Start of outbreaks was measured using a Piecewise regression model.

| City | City census* | ZIP Code population* | Outbreak Start WW | Outbreak Start Cases | $\Delta$ Outbreak Start (Day) | Fold change in SARS-CoV-2 |
| --- | --- | --- | --- | --- | --- | --- |
| SC | 25,435 | 26,739 | 2022-01-04 | 2022-01-11 | 7 | 7.4 |
| R1 | 1,030 | 1,701 | 2022-01-06 | 2022-01-13 | 7 | 15.2 |
| R2 | 763+196† | 2,115 | 2022-01-01 | 2022-01-11 | 10 | 53.9 |
| R3 | 890 | 2,015 | 2022-01-14 | 2022-01-16 | 2 | 21.7 |
| R4 | 624 | 1,167 | 2022-01-05 | 2022-01-09 | 4 | 42.5 |
| R5 | 288 | 985 | 2022-01-12 | 2022-01-12 | 0 | 81.2 |
| * Data from the 2020 Decennial Census obtained from https://data.census.gov/ |  |  |  |  |  |  |
| † Wastewater treatment facility collects effluents from a second city. |  |  |  |  |  |  |

Samples were collected three times a week from October 2021 to March 2022. Rural WWTF samples were time-composite samples collected using Teledyne ISCO 3700 Full Size Portable Sampler (Teledyne ISCO, Lincoln, NE, USA) autosamplers or homemade autosamplers constituted of a Sci-Q 323 peristaltic pump (Watson-Marlow, Falmouth, UK) controlled by an Omron H3CR timer (Omron Corporation, Kyoto, Japan) and housed in a cooler box. Sampling frequencies were comprised between 10 and 30 minutes for 24h. Approximately 3L of wastewater was collected, and subsamples were collected at the end of the 24h sampling period and transported within 6h to the laboratory, where samples were kept at 4°C until further processing. Samples from the SC WWTF were collected using a Teledyne ISCO model 3700 autosampler (Teledyne ISCO, Lincoln, NE, USA), with samples collected paced with influent flow. Sampling failed for less than 10% of the total samples sampled. Samples were kept at 4°C until further processed, at most 3 days later.

Confirmed COVID-19 case counts per zip code were obtained from the Idaho Public Health District 2 website (https://idahopublichealth.com/district-2/novel-coronavirus).

### Sample processing for SARS-CoV-2 detection and quantification

The detailed protocols presented below are publicly available on protocol.io (32). In brief, before concentrating the viral fraction of two replicate wastewater fractions through electronegative membrane filtration, each sample was spiked with the Bovilis® Coronavirus (BCoV) (Merck, Kenilworth, NJ, USA) as a process internal control. Subsequently, filters were inserted together with the DNA/RNA Shield™ (Zymo Research, Irvine, CA, USA) into the Lysis Bead tubes from the AllPrep® PowerViral® DNA/RNA Kit (QIAGEN, Inc., Germantown, MD, USA). Lysis was performed on a FastPrep™ (MP Biomedicals, Santa Ana, CA, USA) for 4 cycles of 20 seconds each at 4.5 m/s and the RNA was then extracted as per the kit manufacturer’s protocol on a QIAcube Connect automated extraction instrument (QIAGEN, Inc., Germantown, MD, USA).

SARS-CoV-2 was quantified by dPCR using the QIAcuity Digital PCR System (QIAGEN, Inc., Germantown, MD, USA) using the GT-Digital SARS-CoV-2 Wastewater Surveillance Assay For QIAcuity® (GT Molecular, Fort Collins, CO, USA). Each 40 µl reaction contained 1x of the Qiagen QIAcuity One-Step Viral RT-PCR Kit (QIAGEN, Inc., Germantown, MD, USA), 1x of the GT Molecular N1-N2-BCoV Assay Solution, and 20 µl RNA template. RNA extraction blanks, dPCR non-template controls and positive controls were included in each dPCR run.

### Data processing and analysis

Fluorescent thresholds were manually set based on the fluorescent level of the positive controls. Then we excluded data from samples for which (i) the recovery rate of the internal processing control BCoV was lower than 1%, or (ii) the RNA extraction process control or dPCR negative control were positive and more than 10% of the measured sample concentration.

The date of an outbreak’s start was determined with a piecewise regression model using either the cumulative sum of the copies per day of the N1 target or the cumulative sum of COVID-19 clinically confirmed cases to estimate the breakpoint in a linear dataset. For each city, we subsampled the linear data around the inflection points corresponding to the dates of the main surge of N1 copies or COVID-19 reported cases in early 2022. Then we fitted a linear regression model in R using the “lm” function with cumulative copies of cases as the response (Y) and the date as the predictor (X). Finally, we fitted the piecewise regression model to the original model, estimating a breakpoint around the inflection of the line, using the segmented() function from the segmented package in R (33).

#### SARS-CoV-2 Epidemiological Model

We constructed a compartmental model to approximate the dynamics of the epidemic in each city. Due to the small size of populations in the rural areas, we expected stochastic effects to be important, and opted to use a discrete-time discrete-state Markov process to approximate the spread of the disease. In this model, individuals in the population can be in one of four states: susceptible (*S*), exposed (*E*), infectious (*I*), and removed (*R*). Importantly, in this model exposed individuals have contracted the disease and shed the virus but are not yet infectious. The changes in the compartments are assumed to be binomially distributed: 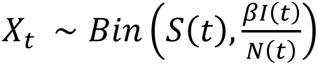 is the number of newly exposed individuals on day 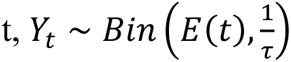 is the number of newly infectious individuals on day t, and 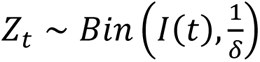 is the number of newly recovered individuals on day t. The parameter *β* is the transmission rate, 𝜏 is the mean incubation period, and 𝛿 is the mean infectious period, and 𝑁(𝑡) = 𝑆(𝑡) + 𝐸(𝑡) + 𝐼(𝑡) + 𝑅(𝑡) is the total population at time t. The discrete-time Markov process is given by:

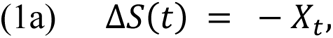

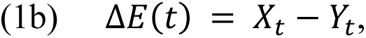

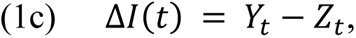

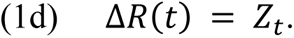

The number of virus particles shed by exposed individuals was assumed to be log-normally distributed (34). However, the log-normal distribution is difficult to work with mathematically, so we approximated the log-normal distribution using the Gamma distribution by matching the first two moments. For simplicity, we assumed that only the individuals in the exposed class 𝐸(𝑡) shed virus in the stools. This assumption is reasonable since it has been shown that the amount of virus shed by a single individual is time-varying, with a peak occurring around symptoms onset (35,36). If there are 𝐸(𝑡) exposed individuals, the amount of virus in the wastewater is a random variable 𝑉(𝑡) with probability density function:

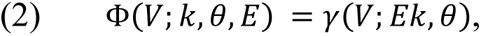

where 𝛾(𝑉; 𝑘, 𝜃) is the probability density function for the gamma distribution with rate 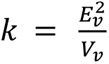 and scale 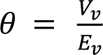 . See Table 2 for parameter values.

**Table 2:** Parameters used in model fitting.

| Parameter | Description | Value | Reference |
| --- | --- | --- | --- |
| $E_v$ | Mean virus copies shed by a single individual | $4.49 \times 10^7$ gc/l | (8) |
| $V_v$ | Variance of virus shed by individuals | $2 \times 10^7$ (gc/l)<sup>2</sup> | (49) |
| $\tau$ | Incubation period | 3 days | (8) |
| $\delta$ | Infectious period | 8 days | (8) |
| $\beta$ | Force of infection | Fit from data | |
| $Q$ | Flow rate of sewershed | Various | Individual treatment plants |
|  | Average faeces produced per day | 128 g | (50) |

We used a sequential Monte Carlo (particle filter) method to fit the collected wastewater data to the stochastic model to the collected wastewater data (Figure 1). In simulations, 50,000 particles (initial conditions) were sampled, using the normalized likelihood distribution for the initial concentrations of virus measured in the wastewater to determine the number of exposed individuals (Figure 1 top row). The initial states of the other classes were sampled uniformly from the remaining population.

**Figure 1:**
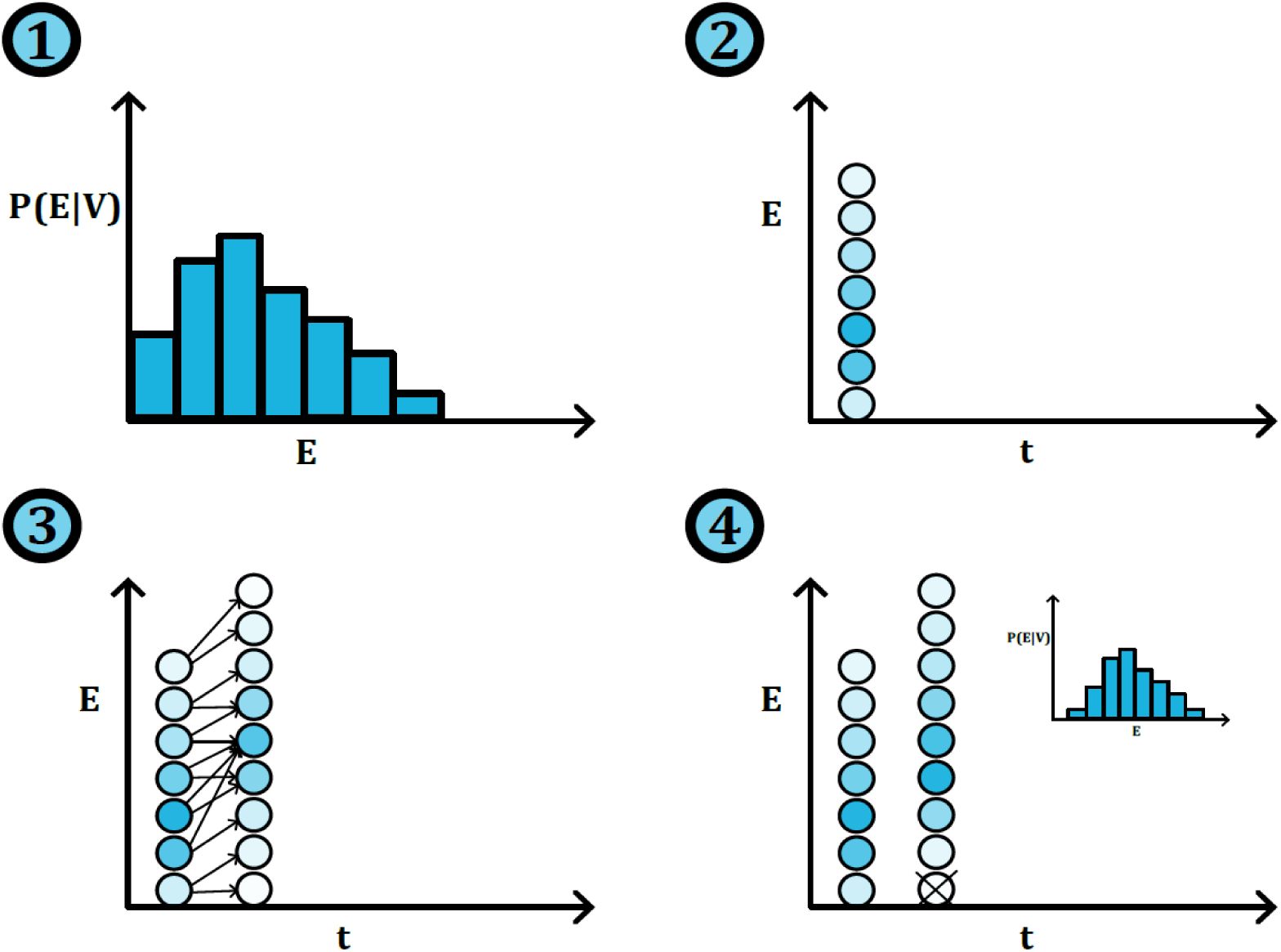
Diagram showing steps of the particle filter method we use to determine the number of active cases from the wastewater titers of SARS-CoV-2. The particle filter is initialized using the first measurement of virus concentration in the wastewater. We generate a distribution of the possible number of infections in the community and sample many (50000) values from this distribution. These values are used as the possible number of exposed individuals (*E*) on day 1 (top right graph). Each of these values also gets a potential number of Susceptible (*S*), Infected (*I*), and Recovered (*R*) individuals. Each set of values (*S,E,I,R*) is called a particle. The darker dots in the diagram signify a higher number of particles with that value of *E*. We apply one step of the stochastic *SEIR* model to each particle to predict the number of infections on the next day (bottom left graph). The measurement of the virus in the wastewater on the next measurement is used to determine which particles are more likely than others. Less likely particles are filtered out using a systematic resampling procedure and replaced with more likely particles (bottom right graph).

Each time step, every particle evolved according to the Markov process in equation 1 (Figure 1 bottom left). On days that we have collected wastewater data, the particles were weighted according to their likelihood (Figure 1 bottom center) and resampled using a systematic sampling method to filter out the least likely particles and reinforce the most likely particles (Figure 1 bottom right).

Data and scripts of this study are available on https://github.com/Tyler-Meadows/wastewater-surveillance.

## Results

### Dynamics of SARS-CoV-2 in rural wastewater vs. clinically confirmed cases

For the period investigated, clinically reported cases revealed that the cities experienced one or two COVID-19 outbreaks, as shown in Figure 2. The first outbreak occurred in late October 2021, but it was not detected in all cities and was relatively small compared to the second outbreak experienced by all cities in early January 2022. This second surge was driven by the Omicron variant, which emerged in the United States in early December 2021 (37).

**Figure 2:**
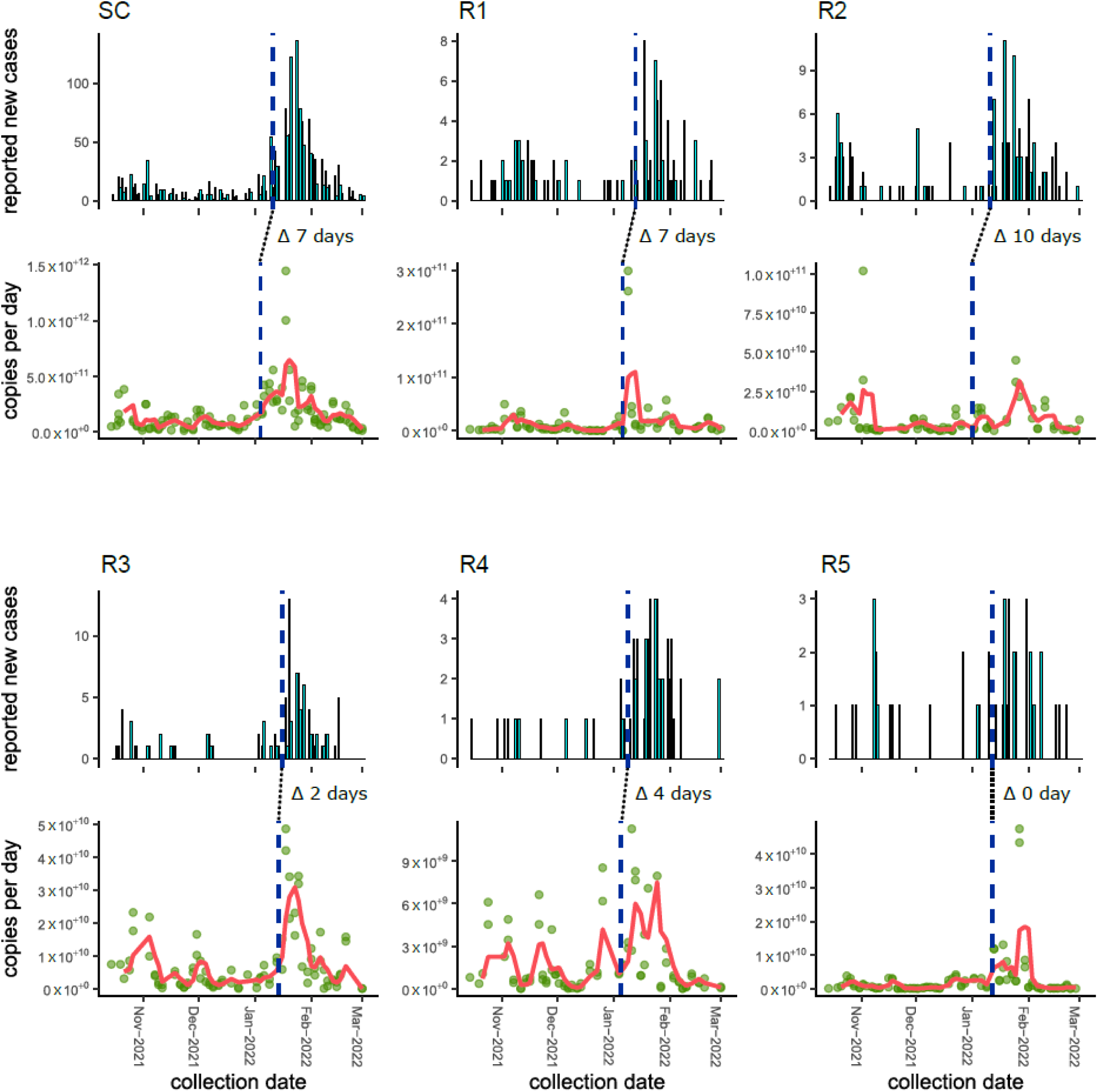
Trend in SARS-CoV-2 in wastewater mirrors the dynamic of the COVID-19 outbreak in rural areas. Each panel represents a city. In each panel, the bar graph shows the time series of the COVID-19 clinically confirmed cases at the specimen collection dates and the second graph shows the measured concentration of SARS-CoV-2 (green dots) with the 7-day moving average (red line). Vertical dash lines represent the estimated start of the outbreak using either the cumulative sum of the copies per day of the N1 target or the cumulative sum of COVID-19 clinically confirmed cases, determined by the Piecewise regression model. Delta shows the difference of days between predicted dates from wastewater-based detection of SARS-CoV-2 and clinically confirmed COVID-19 cases. Cities are ordered by population size (largest on the top left and smallest on the bottom right).

Examining the collected wastewater data, 293 samples were retained following data processing, with each WWTF yielding 44-58 measurements over a five-month period. The daily load of SARS-CoV-2 present in wastewater varied greatly day-to-day, making interpretation of the real-time spread of COVID-19 challenging. Interestingly, the variability in the order of magnitude tended to be larger as the city population decreased, indicating a greater level of randomness (as shown in Figure 3). Additionally, the difference in variance of daily SARS-CoV-2 quantities between cities was significant, as confirmed statistically using Brown-Forsythe, Levene, Barlett, and Kligner-Killeen tests (test results provided in the Supplementary Material). These results suggest that the fluctuations in daily SARS-CoV-2 measurements in smaller cities are more stochastic than in larger cities.

**Figure 3:**
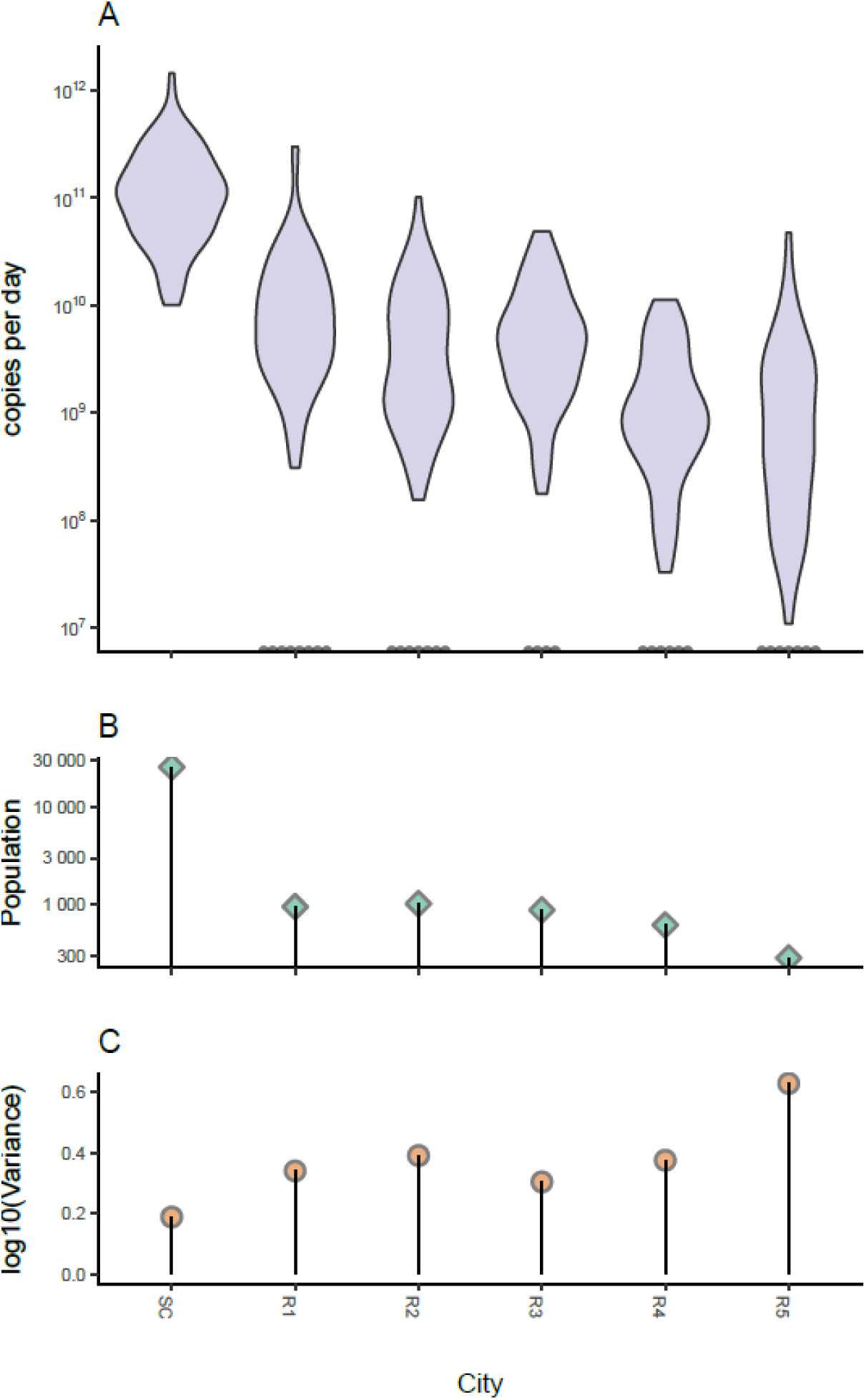
Daily quantities of SARS-CoV-2 tend to be more spread as the city population get smaller. Panel A) shows the distribution of the copies per day of the SARS-CoV-2 on the log scale over the sampling period at each site ordered by city size, detailed in panel B. Note: bin width = 1/30. Dots on the x axis show the samples where N1 was under the detection limit (SC: n = 0, R1: n = 8, R2: n = 8, R3: n = 4, R4: n = 6, R5: n = 7). Panels B) shows the population size of the cities sampled, and C) shows the log scale sample variance measured for each city calculated using the log10 of copies per day of the SARS-CoV-2. This essentially shows that the magnitude of the estimate is less consistent as population size gets smaller (i.e., more stochasticity).

Despite this stochasticity, the Omicron outbreaks resulted in a sharp increase of quantities of the virus collected at the WWTFs above the background levels (Figure 2) by 7- to 81-fold. After estimating the date of the outbreak start (vertical dashed line in Figure 2), we estimated that the SARS-CoV-2 wastewater signal tends to lead the clinically confirmed COVID-19 cases by 0 to 10 days. This supports other retrospective observations, mostly performed in larger cities, that wastewater surveillance could improve and even accelerate the early detection of infectious diseases in rural communities (36,38,39). However, the lead times were variable, and in one case, the wastewater signal was not inferred to precede the clinically reported case data. Other studies have also observed cases where wastewater signal was not preceding clinical testing (38,40).

### Epidemiological model to forecast a COVID-19 outbreak from wastewater detection of the SARS-CoV-2

We used an SEIR-based model (Figure 4A) to investigate whether wastewater-based surveillance of SARS-CoV-2 could enhance the prediction of a COVID-19 outbreak. To test the model’s ability to forecast upcoming trend of cases, we determined if the model could have predicted the Omicron outbreak using exclusively wastewater data. Specifically, we fit the predicted cases using the wastewater measurements up to the onset of the outbreak. This corresponded to the first measurement performed two days after the inflection point defining the start of the outbreak using wastewater data. Then we let the model forecast the upcoming trend in active cases (Figure 4B). When comparing the forecasted COVID-19 cases with the clinically confirmed cases we found that the model successfully captured the clinically active cases in five of the six cities. For the rural cities, the forecasts were within the 95% confidence interval at least up to eight days ahead; corresponding to R4 (Figure 4). While the number of predicted cases tended to be lower than the number of the clinically confirmed cases reported, they were following them closely in two rural cities and in the small city. For those two rural cities (sites R1 and R2), the model accurately predicted the number of active cases more than 14 days ahead. In city SC, the model was able to forecast the number of active cases accurately more than eight days ahead. These results show that in the majority of sewersheds surveyed the model would have confidently predicted the outbreaks even before the COVID-19 reported cases started to increase. Anticipation of the model on the COVID-19 reported cases ranged from 0 days for R5 to 11 days for R2 (see blue dashed line on Figure 4).

**Figure 4:**
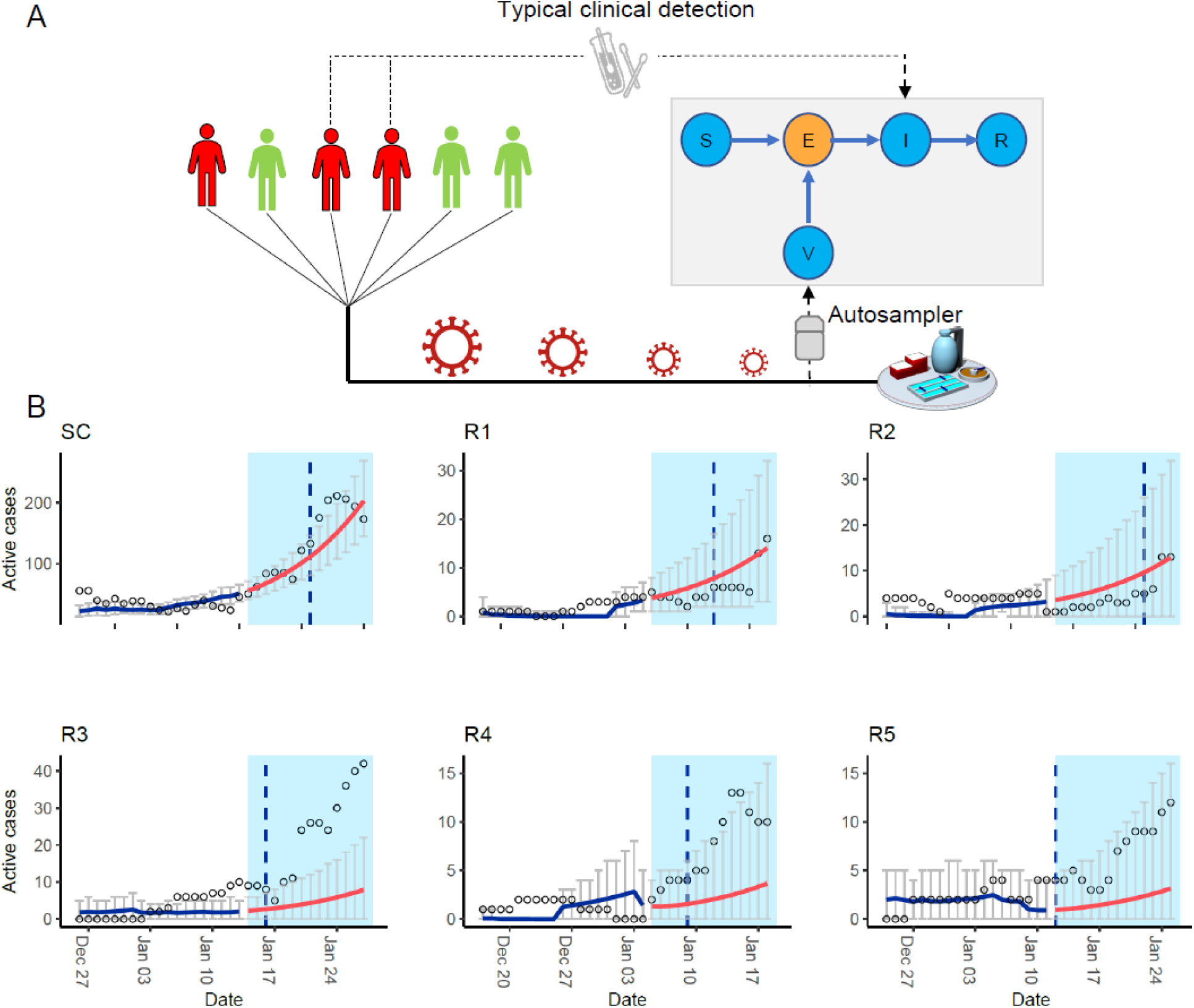
Susceptible-exposed-infectious-recovered model can forecast cases in the early stage of a COVID-19 outbreak. **A)** SEIR model framework depicting a population in green with infected people in red. The SARS-CoV-2 shed by a fraction of the exposed population is measured in the wastewater collected at the WWTF. This titer is integrated into a Susceptible (S), Exposed (E), Infected (I), and Recovered (R) model to estimate the number of exposed individuals E. **B)** Left white side contains known data at the time of the forecast where the blue lines show the fitted predicted active cases from wastewater up to the beginning of the outbreak, and the blue shade shows the data not yet observed at the time of forecast whereas the red lines are active cases forecasted. Vertical dashed lines represent the estimated start of the outbreak based on clinically confirmed COVID-19 cases. Using the wastewater data, the model forecasted the start of the outbreak between 0 to 11 days earlier than the onset of the increase in clinical confirmed cases. 95% confidence intervals are shown by the gray bars. Dots show the active cases determined as the 11-day moving sum of the clinically confirmed cases. Since the mean infectious period from fitting data was 10.88 days, we determined the actual active cases as the 11-day moving sum of new clinically confirmed cases. Breakpoints between fitted and forecast values were chosen to be two days after the start of the outbreak, determined by the Piecewise regression model. Cities are ordered by population size (largest on the top left and smallest on the bottom right).

Since our objective was primarily to test if a SEIR model could predict an outbreak occurring in rural communities, we then focused on the capacity of the model to predict an increase in the trend of active cases. To that end, we used a simulated wastewater dataset to count the number of times the model predicted upward or downward trends in cases correctly (i.e., true positive rate) or incorrectly (i.e., false positive rate). We varied the threshold used to accept predictions to create receiver operating characteristic (ROC) curves (Figure 5). Measured area under the curve (AUC) values presented in Figure 6 reflect the sensitivity and specificity of the forecast for a range of forecasted days. The model tends to predict trends better as the forecast range increases. The ROC curves for predictions made less than a week in advance were significantly lower than those made over nine days (p-values presented in Supplementary Material). After nine days, the AUC medians were above 0.7, and increased to 0.75 at 15 days. These results suggest epidemiologists could rely on these 9 to 15-day forecasts.

**Figure 5:**
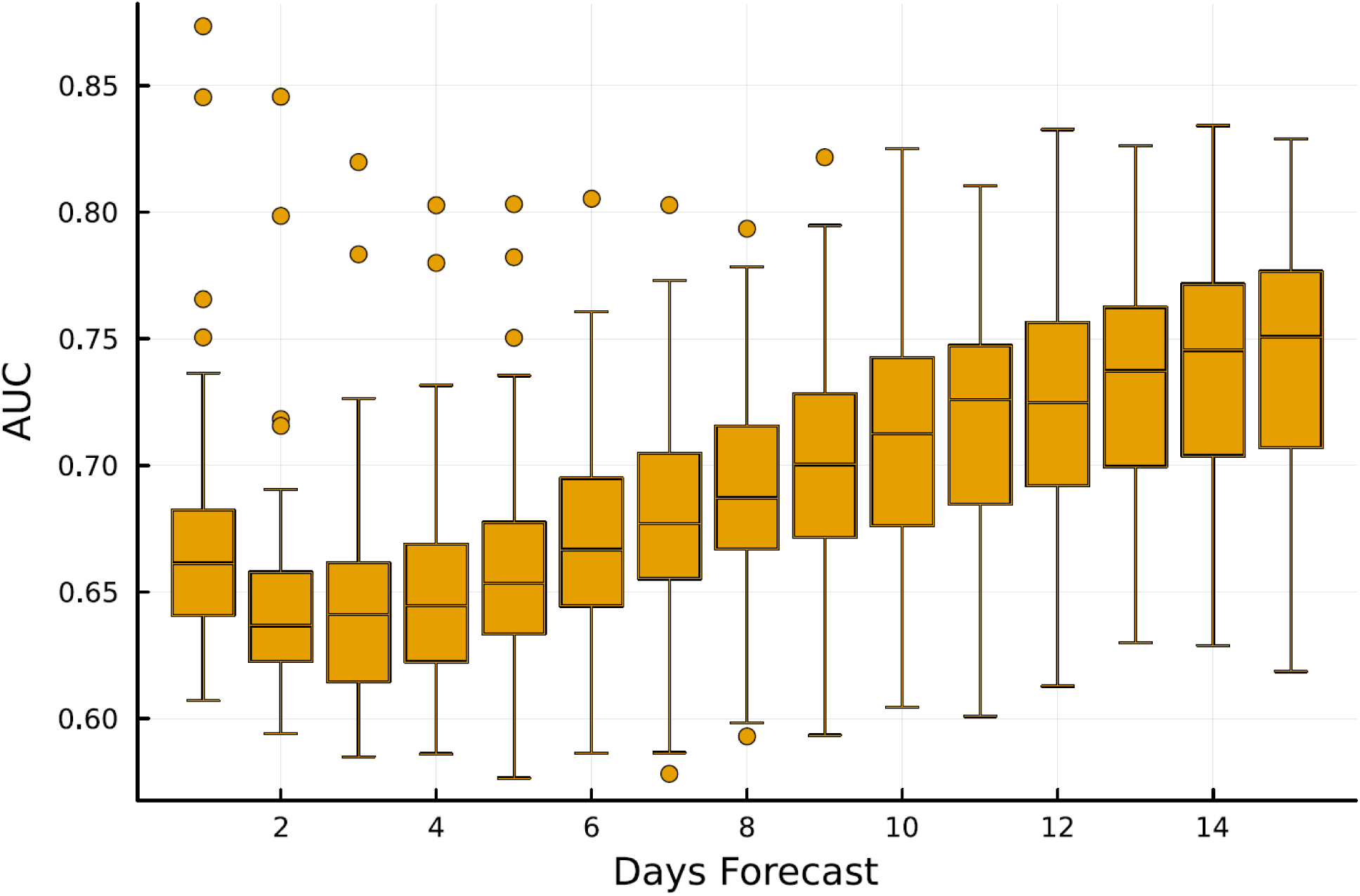
Receiver operating characteristic (ROC) curves for a synthetic data set. A plot showing the number of true increases against false increases for predicted case counts 1,3,5,7,9,11,13, and 15 days beyond the current measurements. A true increase is counted when there was an increase in cases and the model predicted a greater than 𝛼𝛼 probability of an increase. A false increase is counted when there was no increase in cases but the model predicted a greater than 𝛼𝛼 probability of increase.

**Figure 6:**
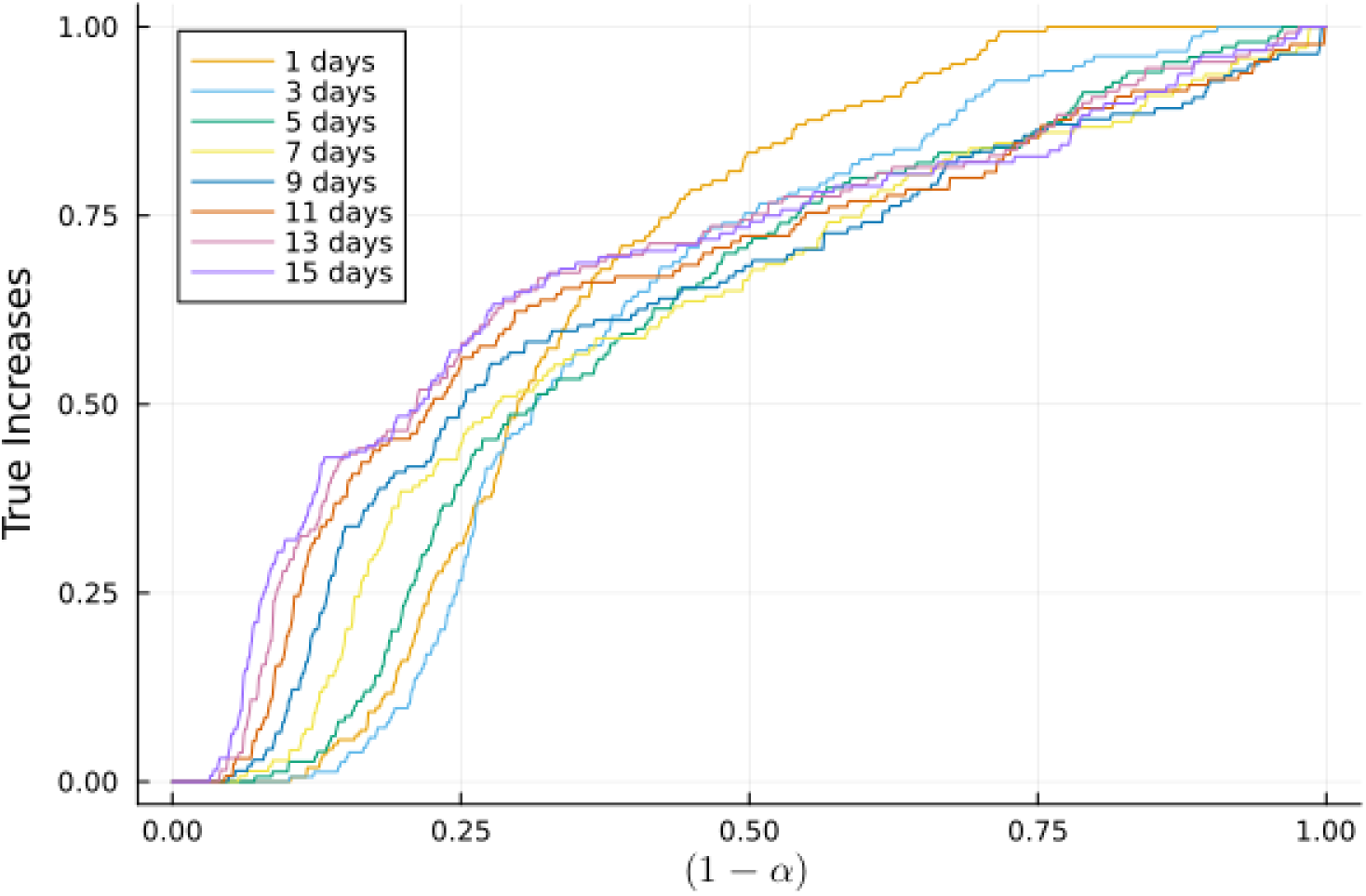
Evaluating SEIR model predictability for an emerging COVID-19 outbreak. Box plot showing the distribution of measured area under the curve (AUC) when computing 50 receiver operating characteristic (ROC) curves when true positive rate is plotted as function of the false positive rate for prediction forecasted from one to 15 days. A random classifier, which represents the outcome if the model randomly picks predictions, has an AUC of 0.5. The further away the curve is from the one of the random classifier, the higher the AUC and the better it illustrates the ability of the model to forecast a trend, with the 1 representing the highest accuracy corresponding to 100% positive rate and 0% false negatives. In general, for a diagnostic test to be able to discriminate patients with and without a disease, the AUC must be above 0.5. Values between 0.7 and 0.8 are considered to be ‘fair’ or acceptable (34,35).

## Discussion

Wastewater-based detection of SARS-CoV-2 has been primarily focused on large urban and metropolitan areas and much less on rural towns. Here we address this gap by having conducted a surveillance effort of the SARS-CoV-2 in the wastewaters of several rural cities of the state of Idaho (USA) since October 2021. Below we discuss the barriers associated with WBE in rural areas and how SEIR modeling could help overcome some of these challenges by forecasting trends in COVID-19 cases based on wastewater-based measurements.

While epidemiologists can examine wastewater data side-by-side with clinical testing to help understand what is happening, many infectious diseases are not reportable, or in the case of COVID-19, at-home self-testing replaced clinical testing (41). For example, influenza and COVID-19 are not reportable diseases in Idaho (USA). Thus, epidemiologists are left with few tools to help them characterize epidemiological trends and forecasts; ultimately only wastewater data may be available to provide insight into disease burden in rural areas.

However, as we experienced, the variability in viral quantities arriving at the rural WWTF per day made the determination of the start of the outbreak in real-time difficult without retrospective statistical analysis. As we observed, the variability in the order of magnitude tended to be larger as the rural sewershed size decreased (42), making interpretation of wastewater-based detection of infectious diseases in rural areas even more complicated for public health. This noise is typically lower in larger sewersheds that are more active over a 24-hr time compared to smaller ones (43).

SEIR epidemiological models offer a framework for epidemiologists to analyze the dynamics of outbreaks using wastewater data (7,8,44–48). Specifically, in our study, we demonstrate that our SEIR model can provide reliable forecasts of when case numbers are trending upwards in rural communities. Sensitivity and specificity assessments of the model in predicting start of the outbreak revealed that the forecasts were more reliable when looking at trends beyond a week, with the best forecast being over a period of nine to 15 days. Similar ranges of short-term forecasts of seven to nine days to predict upcoming cases based on wastewater detection of the SARS-CoV-2 were reported for larger cities using SEIR or SEIR-like approaches (8,46,48). The fact that the model was not performing well under 9 days may, in part, be attributed to the noise of the SARS-CoV-2 quantities in rural wastewater that creates a sawtooth pattern in the trend. Thus, while the trend over a week goes up, there is a chance that some of the points between went down. This led us to hypothesize that the noise in the wastewater data may have made short-term trend predictions less accurate.

While our SEIR model functions well to provide advanced warning of a COVID-19 outbreak, it failed to predict the outbreak peak (Fig. S1) and we could not assess the accuracy of case number predictions. Predicted cases tended to be lower than reported clinical cases for the area, contrasting with other studies on larger sewersheds that have shown that SEIR models typically estimate more COVID-19 cases than the reported number (45,53,54). The underestimation of COVID-19 cases observed in this study may be attributed, in part, to differences between the population of the city sampled and the zip code used (Table 1); the latter corresponds to the clinically recorded cases. When comparing city with zip code census, 39 to 71% of the residents of rural zip codes were not connected to the sewer system of the cities. In rural areas, zip codes often cover a larger geographical area beyond the city limits, which means that comparisons between wastewater data and reported cases should be approached with caution. It is important to note that we did not intend to use the model to predict case numbers; the study was not designed for this purpose. Instead, it was intended to forecast an increase in COVID-19 cases to provide an early warning that could be shared with the community.

Despite the successful application of SEIR models to wastewater surveillance of the SARS-CoV-2, there are some uncertainties on how to connect the model with wastewater data. Some authors have included the cumulative virus titer in the sewershed as a dynamic variable (8) or as a linear combination of other dynamic variables (45). However, this approach can be problematic when measurements are sparse or there are gaps between collection periods – which would be very common in rural WWTFs. Most authors directly connect wastewater measurements to the incidence rate, or prevalence, similar to what we have done herein. In addition, the connection between the disease compartments (the exposed ‘E’ and infected ‘I’ compartments) in the SEIR model and wastewater measurements is not well established. Contribution to viral load in wastewater can tie to the individuals in the ‘I’ compartment (7,8,45) or to the ‘E’ compartment; the SEIR model in this study resulted in better predictions than when connected to ‘I’. This difference could be attributed to the fact that peak virus shedding in stool may occur for a few days around the onset of symptoms (35,36). This means that individuals contributing to the load of SARS-CoV-2 measured in wastewater may be at the transition between ’E’ and ’I’ compartments in the SEIR model. Some researchers have incorporated the results from wastewater measurement in both ‘E’ and ‘I’ (44) while others created a new compartment structure to account for viral shedding dynamic in wastewater (47,48).

Finally, our model was able to successfully forecast the upcoming cases in most of the cities surveyed. In one city this resulted in predicting the outbreak as much as 11 days before reported clinical cases started to increase. However, it failed for one city, suggesting that the disease dynamics do not always follow the model assumptions. This may be because the model does not include potential traveling between cities, which may impact the accuracy of predictions, especially in rural areas where residents often have to commute to work. In the rural cities surveyed between 79.4 and 94.5% of residents work outside their place of residence, versus 27.3% in the small city surveyed (data from U.S. Census Bureau Topic: Commuting – Survey: American Community Survey – 2021, ACS 5-Year Estimates Subject Tables).

In conclusion, our study reveals that wastewater-based epidemiology (WBE) in rural communities and small sewersheds in general is associated with high daily variation in SARS-CoV-2 levels. This variation creates a challenge for epidemiologists who seek to monitor real-time data in rural areas based solely on the raw data. However, our research also shows that the SEIR modeling approach can help to decipher this data and actually predict the start of outbreaks. Our model provides a suitable framework for epidemiologists to analyze the dynamics of outbreaks using wastewater data.

## Acknowledgment

Research reported in this publication was supported by the National Institute of General Medical Sciences of the National Institutes of Health under Award Number P20GM104420. The content is solely the responsibility of the authors and does not necessarily represent the official views of the National Institutes of Health. Data collection for this study was also made possible thanks to the State of Idaho Department of Health and Welfare under grant ## NU50CK000544 funded by the Center for Disease Control and Prevention (CDC) through the Epidemiology and Laboratory Capacity Enhancing Detection Through Coronavirus Release and Relief (CRR) Supplement Funds. The content of this project is solely the responsibility of the authors and does not necessarily represent the official views of the State of Idaho Department of Health and Welfare or the Center for Disease Control and Prevention (CDC).

