## Supplemental material for "Epidemiological model can forecast COVID-19 outbreaks from wastewater-based surveillance in rural communities"

### Variance analysis

We tested if the variance of the total copies of SARS-CoV-2 collected per day at the wastewater treatment facilities, referred to below as "copies_per_day", was different between cities referred to below as "location".

Results from test when testing copies_per_day ~ location

**A) Brown-Forsythe Test (alpha = 0.05)**

```
---------------------------------------------------------------
  data : copies_per_day and location

  statistic  : 69.33715
  num df     : 5
  denom df   : 129.2721
  p.value    : 6.511297e-35

  Result     : Difference is statistically significant.
---------------------------------------------------------------
```

**B) Levene's Test for Homogeneity of Variance (center = median)**

```
       Df F value    Pr(>F)
group   5  27.797 < 2.2e-16 ***
      503
---
Signif. codes:  0 '***' 0.001 '**' 0.01
```

**C) Bartlett test of homogeneity of variances**

```
'*' 0.05 '.' 0.1 ' ' 1

data:  copies_per_day by location
Bartlett's K-squared = 1560, df = 5, p-value < 2.2e-16
```

**D) Fligner-Killeen test of homogeneity of variances**

```
data:  copies_per_day by location
Fligner-Killeen:med chi-squared = 307.03, df = 5, p-value < 2.2e-16
```

**Comparisons of the AUC**

```
Simultaneous Tests for General Linear Hypotheses

Multiple Comparisons of Means: Tukey Contrasts


Fit: glmer(formula = AUC ~ Day + (1 | run_ID), data = AUC_Data, family = gaus
sian(link = make.link("logit")))

Linear Hypotheses:
               Estimate Std. Error z value Pr(>|z|)
2 - 1 == 0   -0.104736   0.023831  -4.395    <0.01 **
3 - 1 == 0   -0.119183   0.023777  -5.013    <0.01 ***
4 - 1 == 0   -0.093857   0.023854  -3.935    <0.01 **
5 - 1 == 0   -0.048525   0.024002  -2.022   0.7852
6 - 1 == 0   -0.004944   0.024168  -0.205   1.0000
7 - 1 == 0    0.046541   0.024376   1.909   0.8484
8 - 1 == 0    0.090456   0.024575   3.681   0.0189 *
9 - 1 == 0    0.135982   0.024787   5.486    <0.01 ***
10 - 1 == 0   0.173965   0.024987   6.962    <0.01 ***
11 - 1 == 0   0.206515   0.025172   8.204    <0.01 ***
12 - 1 == 0   0.245328   0.025392   9.662    <0.01 ***
13 - 1 == 0   0.286689   0.025656  11.174    <0.01 ***
14 - 1 == 0   0.314903   0.025840  12.187    <0.01 ***
15 - 1 == 0   0.331965   0.025964  12.785    <0.01 ***
3 - 2 == 0   -0.014447   0.023379  -0.618   1.0000
4 - 2 == 0    0.010879   0.023458   0.464   1.0000
5 - 2 == 0    0.056211   0.023608   2.381   0.5290
6 - 2 == 0    0.099792   0.023776   4.197    <0.01 **
7 - 2 == 0    0.151276   0.023987   6.307    <0.01 ***
8 - 2 == 0    0.195191   0.024190   8.069    <0.01 ***
9 - 2 == 0    0.240718   0.024405   9.863    <0.01 ***
10 - 2 == 0   0.278701   0.024608  11.326    <0.01 ***
11 - 2 == 0   0.311251   0.024796  12.552    <0.01 ***
12 - 2 == 0   0.350064   0.025019  13.992    <0.01 ***
13 - 2 == 0   0.391424   0.025287  15.479    <0.01 ***
14 - 2 == 0   0.419639   0.025474  16.474    <0.01 ***
15 - 2 == 0   0.436701   0.025599  17.059    <0.01 ***
4 - 3 == 0    0.025326   0.023401   1.082   0.9991
5 - 3 == 0    0.070658   0.023552   3.000   0.1523
6 - 3 == 0    0.114239   0.023720   4.816    <0.01 ***
7 - 3 == 0    0.165723   0.023932   6.925    <0.01 ***
8 - 3 == 0    0.209638   0.024134   8.686    <0.01 ***
9 - 3 == 0    0.255165   0.024351  10.479    <0.01 ***
10 - 3 == 0   0.293148   0.024554  11.939    <0.01 ***
11 - 3 == 0   0.325698   0.024742  13.164    <0.01 ***
12 - 3 == 0   0.364511   0.024965  14.601    <0.01 ***
13 - 3 == 0   0.405871   0.025234  16.084    <0.01 ***
14 - 3 == 0   0.434086   0.025421  17.076    <0.01 ***
15 - 3 == 0   0.451148   0.025547  17.659    <0.01 ***
5 - 4 == 0    0.045332   0.023629   1.918   0.8436
6 - 4 == 0    0.088913   0.023797   3.736   0.0158 *
7 - 4 == 0    0.140398   0.024008   5.848    <0.01 ***
8 - 4 == 0    0.184312   0.024210   7.613    <0.01 ***
9 - 4 == 0    0.229839   0.024426   9.410    <0.01 ***
10 - 4 == 0   0.267822   0.024628  10.875    <0.01 ***
11 - 4 == 0   0.300372   0.024816  12.104    <0.01 ***
12 - 4 == 0   0.339185   0.025038  13.547    <0.01 ***
13 - 4 == 0   0.380545   0.025306  15.038    <0.01 ***
14 - 4 == 0   0.408760   0.025493  16.034    <0.01 ***
15 - 4 == 0   0.425822   0.025619  16.622    <0.01 ***
6 - 5 == 0    0.043581   0.023945   1.820   0.8902
7 - 5 == 0    0.095066   0.024154   3.936    <0.01 **
```

```
8 - 5 == 0    0.138980   0.024355   5.706    <0.01 ***
9 - 5 == 0    0.184507   0.024569   7.510    <0.01 ***
10 - 5 == 0   0.222490   0.024770   8.982    <0.01 ***
11 - 5 == 0   0.255040   0.024957  10.219    <0.01 ***
12 - 5 == 0   0.293853   0.025178  11.671    <0.01 ***
13 - 5 == 0   0.335213   0.025444  13.174    <0.01 ***
14 - 5 == 0   0.363428   0.025630  14.180    <0.01 ***
15 - 5 == 0   0.380490   0.025755  14.773    <0.01 ***
7 - 6 == 0    0.051485   0.024318   2.117   0.7230
8 - 6 == 0    0.095400   0.024517   3.891    <0.01 **
9 - 6 == 0    0.140926   0.024729   5.699    <0.01 ***
10 - 6 == 0   0.178909   0.024929   7.177    <0.01 ***
11 - 6 == 0   0.211459   0.025115   8.420    <0.01 ***
12 - 6 == 0   0.250272   0.025334   9.879    <0.01 ***
13 - 6 == 0   0.291633   0.025599  11.392    <0.01 ***
14 - 6 == 0   0.319847   0.025783  12.405    <0.01 ***
15 - 6 == 0   0.336909   0.025907  13.004    <0.01 ***
8 - 7 == 0    0.043915   0.024721   1.776   0.9075
9 - 7 == 0    0.089442   0.024932   3.587   0.0261 *
10 - 7 == 0   0.127424   0.025130   5.071    <0.01 ***
11 - 7 == 0   0.159974   0.025314   6.320    <0.01 ***
12 - 7 == 0   0.198787   0.025532   7.786    <0.01 ***
13 - 7 == 0   0.240148   0.025794   9.310    <0.01 ***
14 - 7 == 0   0.268363   0.025977  10.331    <0.01 ***
15 - 7 == 0   0.285425   0.026100  10.936    <0.01 ***
9 - 8 == 0    0.045527   0.025126   1.812   0.8937
10 - 8 == 0   0.083510   0.025322   3.298   0.0659 .
11 - 8 == 0   0.116059   0.025505   4.550    <0.01 ***
12 - 8 == 0   0.154872   0.025721   6.021    <0.01 ***
13 - 8 == 0   0.196233   0.025981   7.553    <0.01 ***
14 - 8 == 0   0.224448   0.026163   8.579    <0.01 ***
15 - 8 == 0   0.241510   0.026285   9.188    <0.01 ***
10 - 9 == 0   0.037983   0.025527   1.488   0.9780
11 - 9 == 0   0.070533   0.025709   2.744   0.2771
12 - 9 == 0   0.109346   0.025923   4.218    <0.01 **
13 - 9 == 0   0.150706   0.026181   5.756    <0.01 ***
14 - 9 == 0   0.178921   0.026361   6.787    <0.01 ***
15 - 9 == 0   0.195983   0.026483   7.400    <0.01 ***
11 - 10 == 0  0.032550   0.025900   1.257   0.9956
12 - 10 == 0  0.071363   0.026113   2.733   0.2832
13 - 10 == 0  0.112723   0.026369   4.275    <0.01 **
14 - 10 == 0  0.140938   0.026548   5.309    <0.01 ***
15 - 10 == 0  0.158000   0.026668   5.925    <0.01 ***
12 - 11 == 0  0.038813   0.026289   1.476   0.9793
13 - 11 == 0  0.080174   0.026544   3.020   0.1438
14 - 11 == 0  0.108388   0.026721   4.056    <0.01 **
15 - 11 == 0  0.125450   0.026840   4.674    <0.01 ***
13 - 12 == 0  0.041361   0.026751   1.546   0.9694
14 - 12 == 0  0.069575   0.026927   2.584   0.3798
15 - 12 == 0  0.086637   0.027046   3.203   0.0866 .
14 - 13 == 0  0.028215   0.027175   1.038   0.9994
15 - 13 == 0  0.045277   0.027293   1.659   0.9448
15 - 14 == 0  0.017062   0.027465   0.621   1.0000
---
Signif. codes:  0 ‘***’ 0.001 ‘**’ 0.01 ‘*’ 0.05 ‘.’ 0.1 ‘ ’ 1
(Adjusted p values reported -- single-step method)
```

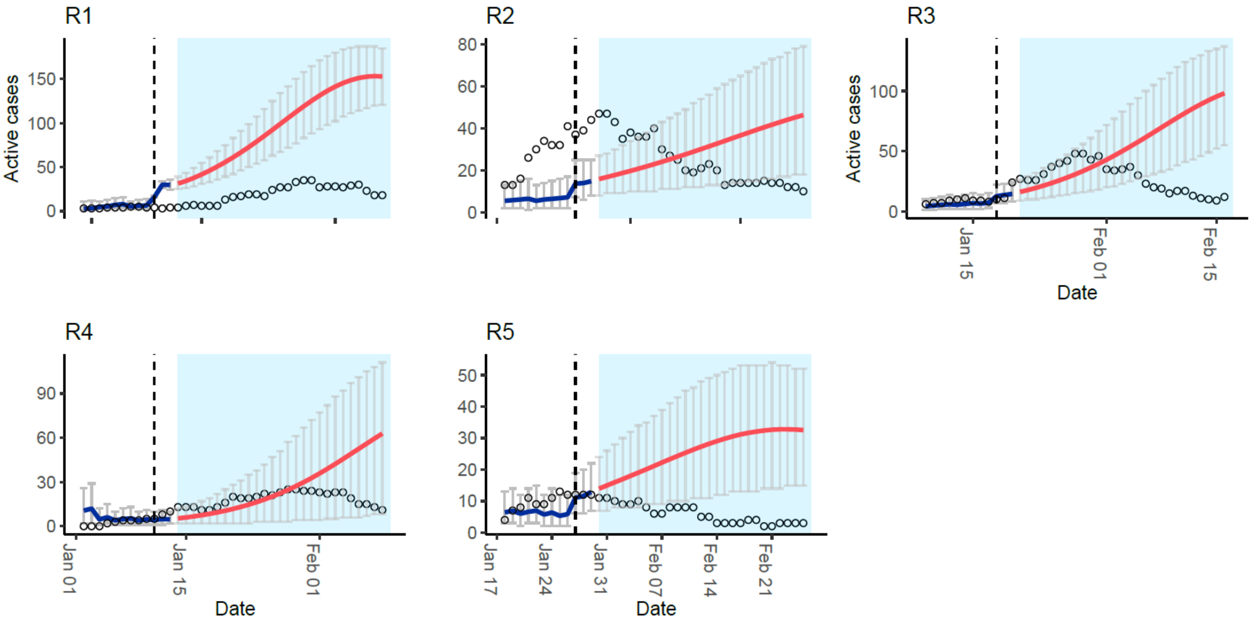


**Figure S1: Outbreak peak forecast.** The red line in the graph represents the predicted number of active cases of SARS-CoV-2 in wastewater. This prediction was made at a time when the virus levels in wastewater were starting to decrease and corresponded to two days after the peak of SARS-CoV-2 in wastewater, which is indicated by the vertical dashed lines. The blue lines in the graph show the estimated number of active cases based on the wastewater data, and the blue shade represents the data that was not yet observed at the time of the forecast. Based on the wastewater data the model failed to predict the outbreak peak accurately. The gray bars indicate the 95% confidence intervals. The dots in the graph represent the number of active cases, which were determined using a moving sum of the clinically confirmed cases over an 11-day period.
